# Significant Relaxation of SARS-CoV-2-Targeted Non-Pharmaceutical Interventions Will Result in Profound Mortality: A New York State Modelling Study

**DOI:** 10.1101/2020.05.08.20095505

**Authors:** Benjamin U. Hoffman

## Abstract

Severe acute respiratory syndrome-coronavirus 2 (SARS-CoV-2) is the most significant global health crisis of the 21^st^ century. The aim of this study was to develop a model to estimate the effect of undocumented infections, seasonal infectivity, immunity, and non-pharmaceutical interventions (NPIs), such as social distancing, on the transmission, morbidity, and mortality of SARS-CoV-2 in New York State (NYS). Simulations revealed dramatic infectivity driven by undocumented infections, and a peak basic reproductive number in NYS of 5.7. NPIs have been effective, and relaxation >50% will result in tens-of-thousands more deaths. Endemic infection is likely to occur in the absence of profound sustained immunity. As a result, until an effective vaccine or other effective pharmaceutical intervention is developed, it will be critical to not reduce NPIs >50% below current levels. This study establishes fundamental characteristics of SARS-CoV-2 transmission, which can help policymakers navigate combating this virus in the coming years.

## Introduction

The global pandemic of severe acute respiratory syndrome-coronavirus 2 (SARS-CoV-2) has emerged as the most significant global health crisis of the 21^st^ century. Since SARS-CoV-2 surfaced in the city of Wuhan, Hubei, China in December of 2019, the virus has caused significant global morbidity and mortality with over 3 million cases and 235,000 deaths worldwide [1-3]. By late April 2020, the United States had accumulated the most cases of SARS-CoV-2 globally, with New York State (NYS) emerging as the epicenter [3]. Local and federal governments have attempted to restrict the spread of SARS-CoV-2 through mandating social distancing, suspending non-essential services, and increasing the testing capacity of health systems [4]. Key unanswered questions are: How effective are these measures at diminishing the active pandemic, and how long might they need to last?

To answer these key questions, the transmission dynamics of SARS-CoV-2 were modelled with a multi-compartment transmission network. This model is capable of simulating the implementation of non-pharmaceutical interventions (NPIs) such as social distancing, contact tracing, and isolation, as well as essential host-pathogen characteristics such as undocumented infection, immunity, and temperature- and humidity-dependent infectivity. For simplicity, NPIs were grouped into a single variable henceforth referred to as “social distancing.” Undocumented infections were defined as individuals infected with SARS-CoV-2, but either due to failure of testing or having been asymptomatic, never undergo quarantine and continue to infect others. The implementation of these factors in a dynamic model of SARS-CoV-2 is essential to not only understanding the fundamental epidemiological dynamics of the current SARS-CoV-2 outbreak, but also to reveal what might drive endemic infection [5, 6].

This study confirms that SARS-CoV-2 is a highly infectious pathogen, whose transmission dynamics are driven by large numbers of undetected undocumented infections. Simulations of relaxed social distancing measures during the summer of 2020 in NYS indicate that modest relaxation is acceptable, but reveal that reduction >50% will dramatically increase transmission and mortality. Projection through 2021 predicts a second outbreak of SARS-CoV-2 in NYS during the winter months, with tens-of-thousands additional deaths if social distancing is not resumed. Lastly, only durable sustained immunity to SARS-CoV-2 will prevent establishment of this virus as an endemic pathogen. Together, these results reveal key characteristics of SARS-CoV-2 transmission dynamics, which are essential to mitigating future morbidity and mortality.

## Results

### Undocumented infections drive SARS-CoV-2 transmission

Undocumented infections represent an increasingly important factor in the spread of SARS-CoV-2. The number of undocumented, but infectious, individuals is a critical variable in modulating the infectivity of respiratory viruses [6, 7]. Recent studies suggest that between 40-95% of all SARS-CoV-2 cases may be undocumented [8, 9]. Serological data from NYS indicate that ~10-20% of the affected population have detectable antibodies to SARS-CoV-2, even though only ~2% have tested positive thus far [10]. Based on these studies, an undocumented infection rate of 75% was selected for this modelling work. An 11-compartment model was developed to comprehensively model SARS-CoV-2 transmission dynamics (see *Methods* and *Supplementary Methods*, **Fig S1**). The model was optimized to SARS-CoV-2 case data from New York State with the particle swarm algorithm, and confidence intervals were estimated by introducing lognormal gaussian noise to the source data and randomly sampling initial parameter estimates (see *Methods*; **Tables S1-2**) [11, 12]. Sensitivity analysis revealed the model was most sensitive to variations of α, which represents the average number of contacts per individual per day (see *Methods;* **Figures S2-3**). Comparison of the model to NYS case-data revealed a good fit (see *Methods;* **Figure S4**).

This model represents an important advancement over recent studies as it incorporates the effect of undocumented infections, seasonal variability, and sustained immunity on SARS-CoV-2 transmission dynamics in NYS (**Figs. 1A-C, S5**) [5, 13-17]. Projecting through September 1^st^, 2020, if the social distancing measures implemented on March 22^nd^, 2020 are not relaxed, the SARS-CoV-2 outbreak will plateau by mid-July 2020: ~1.53 million people in NYS will be infected (undocumented infected+ confirmed symptomatic infected), with ~95,000 total hospitalizations, and ~28,000 deaths (**Fig. 1D-F**) [18]. The maximum basic reproductive number (*R_0_*), which describes the infectivity of a pathogen, over this period was 5.7, significantly greater than the *R*_0_ reported by other published models of SARS-CoV-2 (*R_0_ =* 1.6-3.0) [13, 14, 19]. To account for the rapid growth in testing availability and implementation over the initial period of the outbreak, the maximum basic reproductive number was estimated after the daily rate of change in the ratio of positive tests to confirmed tests was negative for 7 consecutive days (see *Supplementary Methods*). These data demonstrate that the infectious potential of SARS-CoV-2 has thus far been dramatically underestimated, and reveal transmission dynamics that are driven by vast numbers of undocumented infections.

**Figure 1.**
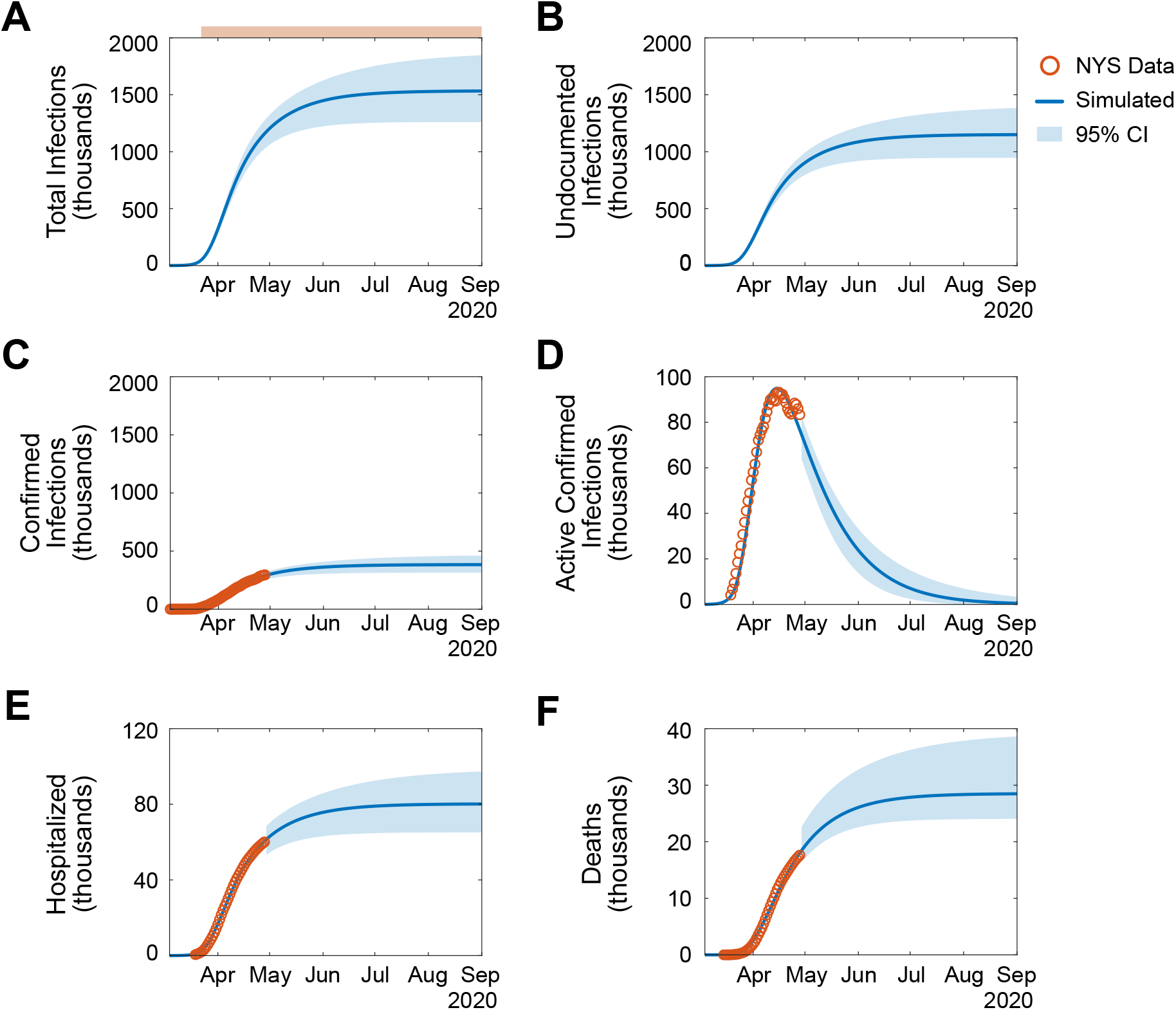
Undocumented infections drive SARS-CoV-2 transmission. **(A-F)**. Simulation of SARS-CoV-2 transmission dynamics through September 1, 2020. Social distancing signified as in (A) top: pink, social distancing initiated on March 22^nd^, 2020. Orange circles, NYS SARS-CoV-2 data (see *Methods*). Blue line, simulated projection. Light blue box, 95% confidence interval. **A**. Cumulative total infections (undocumented infections + confirmed symptomatic infections). **B**. Cumulative undocumented infections. **C**. Cumulative confirmed symptomatic infections. **D**. Active confirmed infections. **E**. Cumulative hospitalizations. **F**. Cumulative deaths. See also, *Figures S2-5*.

### Reduction of social distancing by >50% will result in dramatic mortality

Next, the short-term effects of relaxed social distancing measures in NYS were examined. To do so, a rapid decline in social distancing to a steady-state level over time was simulated until September 1^st^, 2020 (**Fig. 2A-C**, **S6**). Moderate reduction of social distancing (≤50%), independent of the time at which the relaxation was initiated, resulted in minimal differences in total deaths by September 1^st^, 2020 (total deaths: 30%, ~30,000; 50%, ~32,000; **Fig. 2D**). Next, the instantaneous effective reproductive number (*R*(*t*)) was analyzed to quantify the magnitude of social distancing reduction required to initiate a second outbreak of SARS-CoV-2. *R*(*t*) differs from *R_0_* as it describes the fraction of the population that is susceptible to infection as a function of time [20]. Analysis of *R*(*t*) enables temporal estimation of the potential for an outbreak in a population where not all individuals are susceptible: where *R*(*t*) < 1 results in decreasing infections, and *R*(*t*) > 1 results in increasing infections. Only simulations where social distancing was relaxed >50% resulted in *R*(*t*) > 1 at any time before September 1^st^, 2020 (**Fig. 2E-F)** [21]. Simulation of reductions > 50% resulted in the rapid development of a second wave of SARS-CoV-2 cases, with a dramatic increase in deaths by September 1^st^, 2020 (25% vs 75% reduction: ~30,000 vs. ~71,000 deaths; **Fig. 2G-H**). Together, these data indicate that it may be safe to immediately reduce social distancing measures moderately (by up to 50%) in NYS until September 1 ^st^, 2020. However, without effective pharmaceutical interventions policy makers must practice extreme caution in not pursuing increased or continued relaxation, which are predicted to have dangerous outcomes.

Multiple organizations, including the White House, the Centers for Disease Control and Prevention (CDC), and the NYS Department of Health (NYSDOH) have proposed a phased model for relaxing social distancing [22, 23]. These models employ a “wait-and-see” approach to lessening restrictions in order to prevent increased transmission of SARS-CoV-2. To test if a phased approach was superior to a one-time relaxation of social distancing, serial social distancing relaxations, each with a duration of 14 days, were simulated at various time points (**Fig. S7**). For the majority of relaxation magnitudes, one-time relaxation was comparable to phased relaxation; however, phased relaxation was superior for large magnitude reductions in social distancing (one-time 75% vs. phased 75%: ~71,000 vs ~40,000 deaths by September 1^st^, 2020). Together, these simulations demonstrate that phased relaxation should be employed if aggressive social distancing reductions are pursued in NYS.

**Figure 2.**
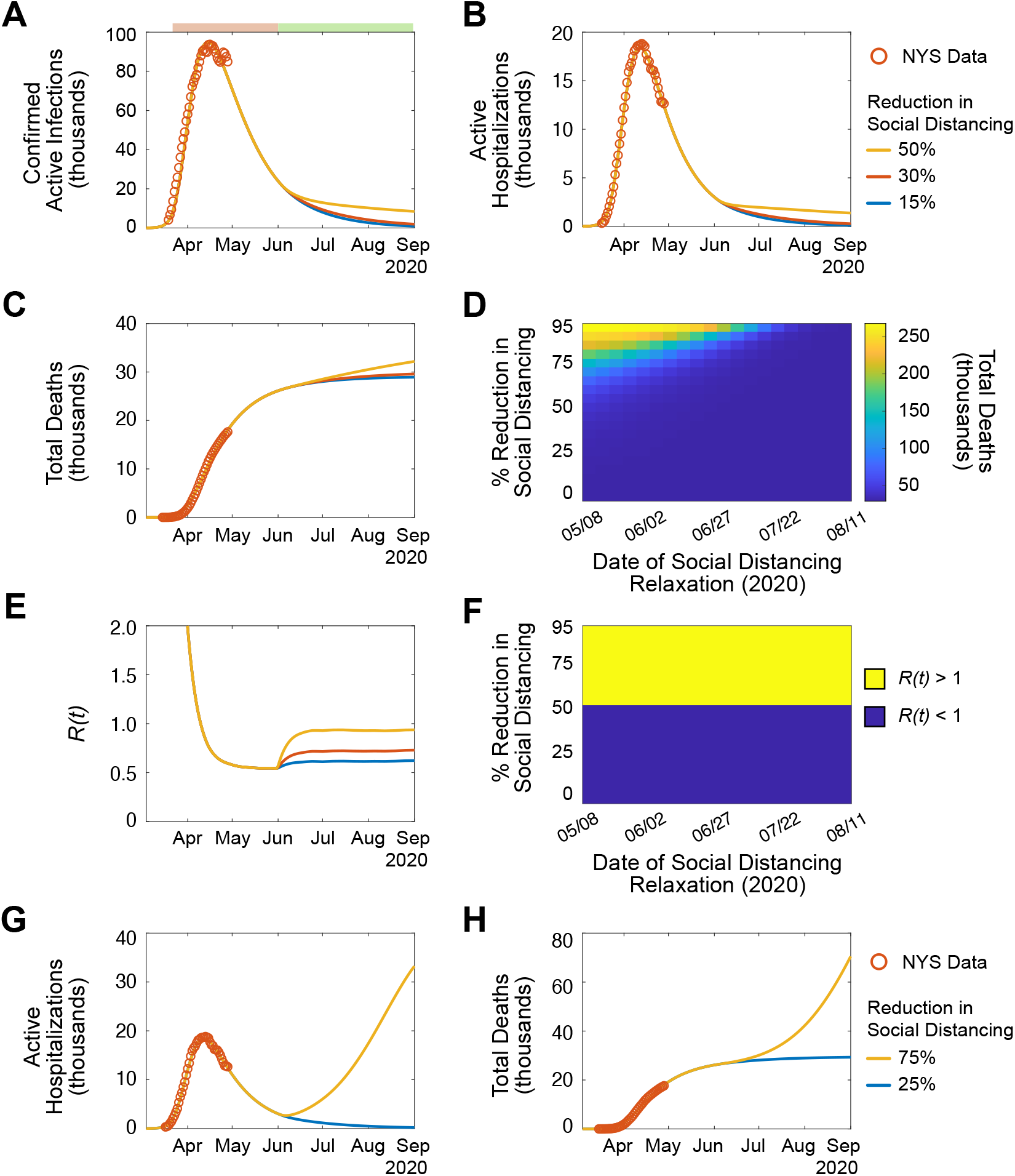
Reduction of social distancing by >50% will result in dramatic mortality. **(A-F)**. Simulation of SARS-CoV-2 transmission dynamics in the presence of social distancing measures through September 1, 2020. Periods of social distancing signified as in (A) top: pink, increased social distancing; green, relaxed social distancing. Orange circles, NYS SARS-CoV-2 data. Lines, simulated projection of reduced social distancing starting June 1, 2020 (yellow, 50% reduction; red, 30% reduction; blue, 15% reduction). **A**. Active confirmed infections. **B**. Active hospitalizations. **C**. Cumulative deaths. **D**. Heatmap displaying the effect of social distancing magnitude and date of reduction on the number of cumulative deaths. **E**. *R*(*t*). **F**. Categorical heatmap displaying the effect of social distancing magnitude and date of reduction on *R*(*t*) *>* 1 (yellow, *R*(*t*) > 1; blue *R*(*t*) < 1). (**G-H**). Simulation of extreme reduction of social distancing on June 1, 2020 (yellow, 75%; blue, 25%). **G**. Active hospitalizations. **H**. Cumulative deaths. See also, *Figures S6-7*.

### Recurrent outbreak of SARS-CoV-2 in NYS in early 2021

Given the social and economic toll and magnitude of the current SARS-CoV-2 pandemic, a key question is: will there be a recurrent outbreak in the next 1.5 years? To answer this question, the course of SARS-CoV-2 was simulated through September 1^st^, 2021 (**Fig. 3A-C; Fig. S8**). Indeed, a severe SARS-CoV-2 outbreak is predicted to occur through the winter months of 2020/21. These results reveal the significant effect of temperature- and humidity-dependent infectivity of SARS-CoV-2, which were modelled based on the transmission dynamics of other non-SARS-CoV-2 human coronaviruses (HCoVs; see *Methods*). The magnitude of the outbreak depends on the resumption of social distancing measures, where stronger measures implemented early result in significantly reduced morbidity and mortality through September 1^st^, 2021 (% social distancing, cumulative deaths: 100%, ~58,000; 75%, ~92,000; 50%, ~154,000; **Fig. 3D**). However, analysis of the *R*(*t*) indicates that the potential for a second outbreak is extremely high, where no combinations of social distancing strength and implementation date were successful in preventing *R*(*t*) > 1 (**Figs. 3E, F**). Together, these data predict a significant second outbreak in early 2021 that can be mitigated, but not avoided entirely, through the resumption of strong social distancing measures.

**Figure 3.**
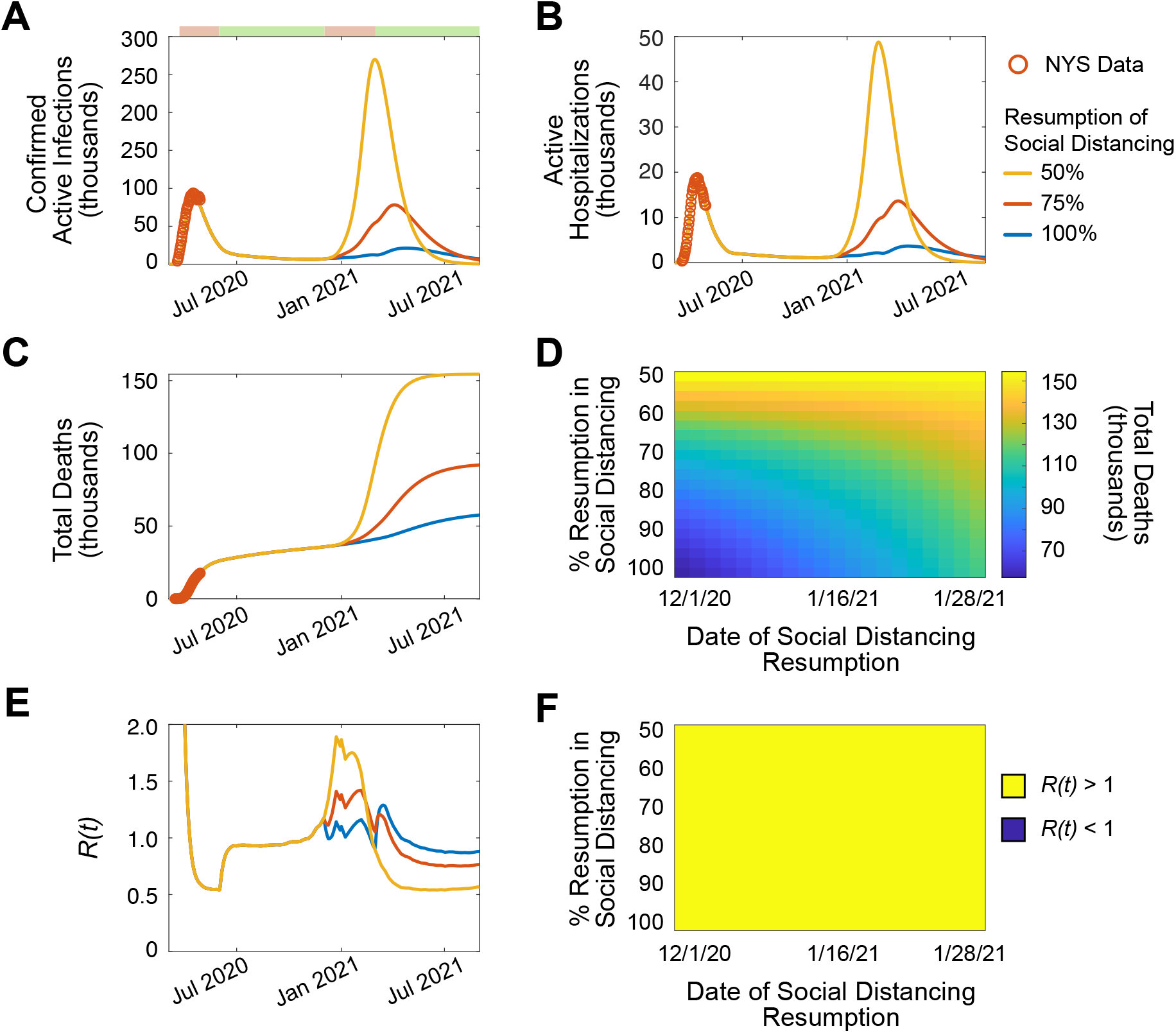
Recurrent outbreak of SARS-CoV-2 in NYS in early 2021. (**A-F**). Simulation of SARS-CoV-2 transmission dynamics in the presence of social distancing measures through September 1, 2021. Simulation assumes 50% social distancing reduction on June 1, 2020. Periods of social distancing signified as in (A) top: pink, increased social distancing; green, relaxed social distancing. Orange circles, NYS SARS-CoV-2 data. Lines, simulated projection with resumption of social distancing on December 1, 2020 to Feb 28, 2021, and relaxation after March 1, 2020. **A**. Active confirmed infections. **B**. Active hospitalizations. **C**. Cumulative deaths. **D**. Heatmap displaying the effect of social distancing magnitude and date of resumption on the number of cumulative deaths. **E**. *R*(*t*). ***F***. Categorical heatmap displaying the effect of social distancing magnitude and date of resumption on *R*(*t*) *>* 1 (yellow, *R*(*t*) > 1; blue *R*(*t*) < 1). See also, *Figure S8*.

### The endemic potential of SARS-CoV-2 depends on immunity

Respiratory viruses, including non-SARS-CoV-2 HCoVs and influenza are endemic across the world, in part due to the seasonal variability of their infectivity and incomplete sustained immunity [24, 25]. Thus, it is essential to understand what factors might drive endemic infection of SARS-CoV-2 in NYS. To do so, the transmission dynamics of SARS-CoV-2 were simulated through September 1^st^, 2025 (**Figs. 4A-B; Fig. S9**). Indeed, SARS-CoV-2 emerged as an endemic pathogen in NYS despite resumption of social distancing for 3 months in the winter of 2020/21. Next, the effects of differential immune parameters (% infected individuals that become immune, and duration of immunity) on cumulative deaths and endemic potential were analyzed (**Figs. 4C-D**). Only parameters conferring large proportions of infected individuals (>70%) with long-term sustained immunity (>15 years) resulted in eventual elimination of annual SARS-CoV-2 infection over 5 years. These data demonstrate that the potential for SARS-CoV-2 to become an endemic pathogen in NYS directly depends on the quality of immunity that individuals develop. As a result, the development of an efficacious vaccine will be essential to prevent endemic SARS-CoV-2 infection in NYS.

**Figure 4.**
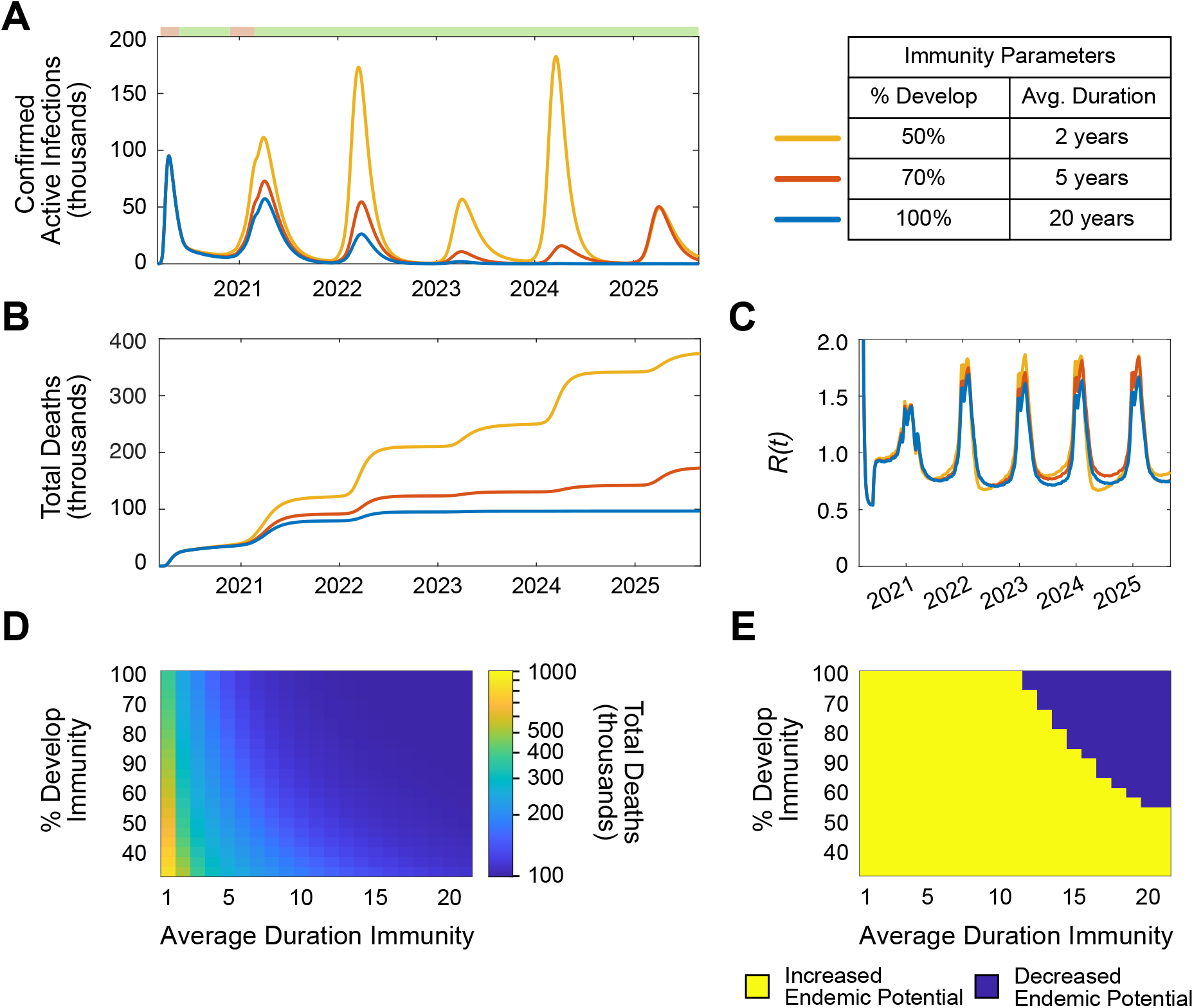
The endemic potential of SARS-CoV-2 depends on immunity. (**A-D**). Simulation of SARS-CoV-2 transmission dynamics in the presence of social distancing measures through September 1, 2030. The following social distancing parameters were assumed: 50% from June 1 to October 31, 2020; 75% from November 1, 2020 to March 1, 2021; 50% after March 1, 2021. Periods of social distancing signified as in (A) top: pink, increased social distancing; green, relaxed social distancing. (**A-B**). Simulation varying the % of individuals who develop immunity and the average duration of immunity. **A**. Active confirmed infections. **B**. Cumulative deaths. **C**. Heatmap displaying the effect of immunity parameters on cumulative deaths through September 1, 2025. **D**. Heatmap displaying the effect of immunity parameters on endemic potential. Endemic potential defined as the presence annual recurrence (average total annual infections > 20,000) from September 1, 2023-2025. See also, *Figure S9*.

## Discussion

As the global health community continues to grapple with issues caused by the growing spread of SARS-CoV-2, it is essential to develop insights into the complex transmission dynamics of this pathogen. Mathematical models can be extremely useful to reveal epidemiological characteristics of infections [26]. Here, an epidemiological model was developed to predict the spread, morbidity, and mortality of SARS-CoV-2 in NYS. This model is designed to describe key features of the virus, such as the effect of undocumented infections on transmission, the development of sustained immunity to the virus, change in infectivity with weather, and the effect of non-pharmaceutical interventions to mitigate the outbreak. This study reveals that dynamic social distancing measures are essential and critical to controlling the spread of SARS-CoV-2, and that in the absence of development of profound immunity caused by infection or the development of an efficacious vaccine, the virus is very likely to become endemic.

One important finding of this study is that vast undocumented infections fueled the rapid spread of SARS-CoV-2 in NYS. During the recent NYS outbreak, the model estimated a peak *R_0_* of 5.7, reaching a baseline of 4.4 by July 2020, far higher than reported in other publications (**Figure S5**): which estimated 2.2 in the United States, and 2.2-2.6 in Wuhan and other regions of China [5, 13, 14, 27]. The magnitude of the model’s estimated *R_0_* reflects the vast number of predicted undocumented infections in NYS at the start of the outbreak. Based on serological studies, a 75% undocumented infection rate was implemented in this study (see *Methods*) [8-10]. As a result, the model projects ~688,000 undocumented infections by April 28^th^, 2020, despite the NYSDOH reporting only ~295,000 confirmed infections. Together, these undocumented infections explain the exponential growth of the outbreak in NYS: undocumented infected individuals were unaware of having contracted infection with SARS-CoV-2, and rather than entering quarantine, continued to expose contacts with the virus.

The effect of social distancing on the course of SARS-CoV-2 in NYS reveals that NPIs are effective in reducing the spread of the virus; however, to prevent the development of large magnitude outbreaks capable of overwhelming health-care systems, they will likely have to persist for multiple years. These measures can be reduced modestly to about 30% of current levels without having a significant impact on SARS-CoV-2-related morbidity and mortality. However, if they are reduced more than 50%, *R*(*t*) will become >1, and morbidity and mortality will dramatically increase. NPIs will need to rely not only on social distancing, but also on improved testing, isolation, and contact tracing. Indeed, intensive testing and case-based interventions have become essential to control efforts in countries across the globe [28-30]. To simulate this long-term steady state of improved NPIs, the social distancing variable was never allowed to reach 0% in long-term simulations (**Figs. 3, 4, S8, S9**).

Although this model employs data obtained from the SARS-CoV-2 outbreak in NYS, the conclusions made herein can be extrapolated to cities globally. To facilitate this extrapolation, the infectivity within the model is proportional to population density (**ρ**; **Table S1**; see *Methods*), as rapid spread of SARS-CoV-2 likely depends on high population densities [31]. Thus, the model can be applied to any city, region, or country by modifying the population density variable. For example, while NYS experienced rapid SARS-CoV-2 transmission, the outbreak in the San Francisco Bay Area was significantly smaller. This was in part due to the earlier shelter in place order (March 16^th^, 2020 vs. March 22^nd^, 2020 in NYS), but may also reflect a less dense population [18, 32]. The San Francisco Bay area counties (San Francisco, San Mateo, Santa Clara, Alameda, and Marin) together have a population density of 1.809 (thousands per sq. mile), compared to the 2.726 of the New York counties in this study [33]. As a result, the San Francisco Bay Area would be predicted to have lower SARS-CoV-2 infectivity. Future implementations of this model will be important to understand the differential transmission dynamics of SARS-CoV-2 in cities across the United States, and the world.

The conclusions of this study build upon the growing body SARS-CoV-2 modelling work by incorporating the effects of NPIs, undocumented infection, seasonal infectivity, and sustained immunity into a single model of NYS transmission. Other studies have modelled these variables in part, and/or in other regions across the word. For example, a number of studies have concluded that NPIs are essential to mitigating SARS-CoV-2 transmission [13-17, 19]. Additionally, a recent study revealed that substantial undocumented infections were essential to the rapid dissemination of SARS-CoV-2 in China [5]. Seasonal infectivity has been analyzed by other models, which together also predict seasonal establishment of endemic SARS-CoV-2 transmission [14, 34]. Here, the effects of these variables were all incorporated into a single model; thereby enabling dynamic analysis of their effects on short- and long-term transmission of SARS-CoV-2.

This model has a number of limitations. First, the model relies on data obtained from the NYSDOH, including public reports and press conferences. These data likely have errors in their estimations; thus, the model projections may overestimate or underestimate true epidemiological characteristics. Additionally, these data do not account for the relative lack in testing that occurred during the initial period of the NYS outbreak. As a result, the reported confirmed cases likely are under representative, which may influence the model towards higher infectivity. Second, a fixed parameter approach was chosen for a number of variables including average incubation period (γ^−1^), average time to hospitalization (δ^−1^), latency to non-hospitalized quarantine (θ^−1^), and the effective rate of undocumented infections (ν; see *Supplementary Methods;* **Table S1**). The values of these parameters were chosen based on literature; however, in reality they likely change over time. Third, the seasonal variability SARS-CoV-2 transmission was modelled after non-SARS-CoV-2 HCoV seasonal variability. This assumption was conservatively weighted because the sensitivity of SARS-CoV-2 transmission to temperature and humidity is not yet known (see *Methods*); therefore, the true seasonal variability of SARS-CoV-2 may differ from the model’s predictions. Lastly, pharmaceutical interventions, such as vaccines and therapeutic and prophylactic medications were not included in this model. It is unknown when these pharmaceutical interventions will be available, thus, there could be no confidence in any estimation of their emergence. However, these will likely represent the most critical long-term interventions in the global health arsenal to combat endemic SARS-CoV-2. The development of these pharmaceutical interventions will be essential to reduce the ultimate toll of this significant pandemic.

SARS-CoV-2 represents the most significant global health crisis of the 21^st^ century. This modelling study reveals fundamental characteristics of this pathogen, which will inform policymakers combating recurrent SARS-CoV-2 infection in the coming months to years. Simulations reveal that NPIs, such a social distancing, are highly effective in reducing short-term morbidity and mortality, but only sustained immunity can prevent SARS-CoV-2 from emerging as an endemic pathogen. As a result, pharmaceutical interventions, such as highly effective drugs, antibodies, or vaccines will ultimately be necessary to combat SARS-CoV-2 in the future.

## Methods

### Data

SARS-CoV-2 case data for NYS were extracted from publicly reported clinical statistics by the New York State Department of Health, aggregated by the COVID Tracking Project [35, 36]. The data extracted were the total number of confirmed cases, total number of hospitalizations, active hospitalizations, and total number of deaths. Since the overwhelming majority of SARS-CoV-2 cases are concentrated in a limited number of NYS regions, only counties reporting greater than 1000 confirmed cases on (4/20/2020) were included (Queens, Kings, Nassau, Bronx, Suffolk, Westchester, Manhattan, Rockland, Richmond, Orange, Dutchess, Erie, and Monroe). To estimate population density of the affected NYS counties, population and land area estimates were obtained from the United States Census Bureau [33].

Non-SARS-CoV-2 human coronavirus (HCoV) viral testing data was obtained from the CDC National Respiratory and Enteric Virus Surveillance System (NRVESS) [37]. Data were extracted from two databases: viral testing data from 2014-2017 and 2018-2020 [38, 39]. To estimate the incidence of the HCoVs, the weekly percentage of positive tests were multiplied by the weekly population-weighted proportion of physician visits due to influenza-like illness (ILI) reported by the US Outpatient Influenza-like Illness Surveillance Network (ILINet) [38]. This method was based on methods published elsewhere [14, 40]. All data were compiled in Matlab (*Mathworks*) and prepared with custom-built software.

### Transmission model of SARS-CoV-2

The transmission dynamics of SARS-CoV-2 were modelled with a system of ordinary differential equations (ODEs) describing an eleven-compartment susceptible-exposed-infected-recovered-susceptible (SEIRS) model (see *Supplementary Methods*) [41]. The schematic of the model is shown in **Figure S1**. “Confirmed active infections” were calculated as the sum of Q and H. The compartment P represents individuals protected by NPIs, such as social distancing measures. The following coefficients were implemented as functions of time: α(*t*), ζ(*t*), λ(*t*), κ(*t*), and χ(*t*). To model the implementation of social distancing measures, α=0 before social distancing measures were in place, and became a fitted constant after (social distancing initiated in NYS on March 22^nd^, 2020) [18]. The “leak” of individuals from the protection of social distancing was modelled with *ζ*(*t*), where the magnitude of ζ was manipulated in simulations to represent the lessening of social distancing measures. The recovery and death rate of hospitalized patients were modelled with λ(***t***) and κ(***t***) respectively, which were composed of the follow one-phase exponential association and decay equations:

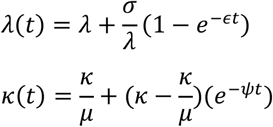

Seasonal variability of SARS-CoV-2 infectiveness was modelled with χ(*t*). To do so, the weekly incidence of HCoVs from 2014-2020 were averaged to create a generalized annual model of HCoVs seasonal variation. It is yet unknown how the infectiveness COVID may vary with weather. Recent studies including retrospective analyses, *in vitro* experiments, and epidemiological models suggest there may be a component of weather-based variability; however, this may be significantly less other respiratory viruses [42-48]. Based on these studies, the seasonal variability of SARS-CoV-2 (*χ*(*t*)) was generated by conservatively weighting the generalized annual model of HCoV seasonal variability by a factor of 0.2.

Undocumented infections represent an increasingly important factor in the spread of SARS-CoV-2. From serological studies, an undocumented rate of 75% was estimated by dividing the total number of confirmed cases by the product of the percent of serologically positive individuals and the population of the most affected NYS counties (see *Methods, Data*), subtracted from one [8-10]. To calculate ν, the following formula was used:

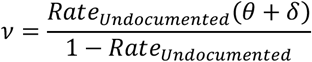

The duration and annual recurrence of SARS-CoV-2 infections depends on the fraction of people who develop protective immunity to SARS-CoV-2, and the duration of immunity. It is currently unclear how individuals will respond to SARS-CoV-2 infection; however, data from similar HCoVs may provide an estimated range. Studies suggest that HCoV-OC43 and HCoV-HKU1 confer protective immunity that lasts less than a year, whereas immunity to SARS-CoV-2 may last much longer [24, 25]. To model immunity, the transition rate from “Recovered” compartments (R_A_, R_Q_, and R_H_) to the “Susceptible” compartment depended on the following relationship:

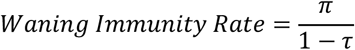

Where π represented the inverse of the duration of immunity, and τ represented the fraction of individuals who do not develop immunity.

Parameter estimation, sensitivity analysis, goodness of fit analysis, and reproductive number proofs are described in *Supplementary Methods*.

### Simulations and data analysis

Simulations and output figures were generated in Matlab (*Mathworks*), with custom-built software. Figures were edited in Adobe Illustrator (*Adobe*).

## Data Availability

The custom-built software used generate this model, along will scripts to create the analyses and data within this manuscript, is freely available at https://github.com/buh2003/BUHoffman_COVID

## Data and software availability

The custom-built software used generate this model, along will scripts to create the analyses and data within this manuscript, is freely available at https://github.com/buh2003/BUHoffman COVID.

## Acknowledgements

Thank you to Dr. Stephen L. Hoffman, Dr. Kim Lee Sim, and Dr. Seth A. Hoffman for helpful discussions and comments on the manuscript; Dr. Steven Shea for essential feedback on preliminary studies; and Mr. Abulhair Saparov for input on the methods and analysis. The author was funded by a NIH MSTP training grant (NIGMS T32GM007367).

